# Synthetic Data Distillation Enables the Extraction of Clinical Information at Scale

**DOI:** 10.1101/2024.09.27.24314517

**Authors:** Elizabeth Geena Woo, Michael C. Burkhart, Emily Alsentzer, Brett K Beaulieu-Jones

**Author notes:** Correspondence: Brett Beaulieu-Jones. Authors contributed equally (Elizabeth Geena Woo, Michael C. Burkhart).

## Abstract

Large-language models (LLMs) have shown promising potential for extracting information from clinical notes. Deploying these models at scale can be challenging due to high computational costs, regulatory constraints, and privacy concerns. To address these challenges, we used synthetic data distillation to fine-tune smaller, open-source LLMs that achieve performance similar to that of larger models, including the teacher model. These smaller models can be run on less expensive local hardware or at a vastly reduced cost in cloud deployments. In this study, we used Llama-3.1-70B-Instruct to generate synthetic training examples in the form of question-answer pairs along with supporting information and model-assigned difficulty scores. These synthetic examples were used to fine-tune the smaller Llama-3.1-8B-Instruct model. We evaluated the performance of these models on an annotated synthetic dataset resembling clinical trial criteria, the i2b2 2018 Clinical Trial Eligibility Challenge, and clinical notes reflecting the clinical trial for apixaban. The fine-tuned models outperformed the 8B-Instruct model on all tasks and in some cases even exceeded the performance of the larger 70B-Instruct model. This work demonstrates the potential of synthetic data distillation to enable more scalable and efficient clinical information extraction, which could be applied toward improving accuracy and efficiency of patient phenotyping and clinical-trial matching.

## Introduction

Research with real-world data typically relies on human-labeled data for training and validation. Though effective, human annotation can be costly, time-consuming, and prone to errors. Recent research suggests that few-shot capabilities of generative large language models (LLMs) can be used to annotate text data with reduced time and cost burden^1–4^. These capabilities of generative LLMs can be applied to information extraction from patient clinical notes. Traditional methods for information extraction include rule-based approaches, which can be limited by low recall due to user-defined rules and variability of medical texts, and supervised machine learning models, which can be limited by lack of labeled training data^5–7^. The zero- and few-shot capabilities of LLMs can enable more flexible and scalable information extraction from clinical notes without the need for extensive manual annotation.

While promising, state of the art LLMs (such as GPT-4^8^) are challenging to deploy in a scalable way in healthcare systems. Many of these models (including those from OpenAI, Anthropic, and Google) are proprietary and come with limited license terms. Concerns about patient privacy and lack of transparency in these proprietary models also lead to some hesitancy in their adoption for healthcare institutions^9^. Additionally, these models can be extremely large and require substantial computational resources (e.g., Llama 405B), limiting their deployment within typical health system IT settings^10^. So far, many of the successful deployments have been through partnerships where industry partners may subsidize cost or provide in-kind contributions in terms of compute and engineering. This may limit the number and type of institutions who are able to participate and the use cases they are able to apply generative AI to. Additionally, setting up these partnerships can require additional administrative lift (e.g., legal negotiation and information security evaluation) compared to performing analyses in existing environments, whether institution-hosted or existing private cloud deployments^11^. Even where solutions have been widely available, such as partnerships for draft inbox responses^12^, the ability to achieve similar performance with smaller models will make customizing models to a specific institution as well as serving inference requests at scale substantially cheaper and less cumbersome.

Challenges in generative AI around scalability necessitate cost-effective and privacy-conscious solutions, which could be addressed through the development of open-source LLMs that can be integrated into existing healthcare system infrastructure. Open-source LLMs historically did not perform as well as their proprietary counterparts^13^ but recent progress has led to very competitive models across most evaluation metrics^14^. Recent efforts have been made to evaluate the capacity of locally deployable LLMs to extract clinical information with low hardware requirements^15^. Synthetic data generation, distillation, and instruction tuning offer an opportunity to close the gap between open-source and proprietary models. Larger models can generate synthetic data that can be used to fine-tune a smaller model for a given task, with the idea that the smaller model could mirror the performance of the larger model for that task. This process, called *distillation*, has been shown to improve performance of these models, particularly when there is less available labeled data such as paired patient-criterion matching annotations for patient-trial matching. It allows researchers to develop models with the potential for wider adoption through reducing computational cost without sacrificing performance.

The ability to extract clinical information at scale from unstructured clinical notes could enhance patient phenotyping, which is important for research and clinical applications. Current phenotyping approaches often rely on structured data such as ICD codes, which are used for billing purposes and may not reflect the nuances of the patient’s condition. This can limit analytical precision and potentially introduce biases when studying research outcomes. Unstructured clinical notes, which contain information including medical, social, and family history that may not be captured by structured data, could offer more granular and reliable insight into patient history, particularly in heterogeneous populations where there can be large differences in disease manifestation and progression^16,17^. LLMs can perform zero-shot information extraction from notes that improve phenotyping accuracy over the use of ICD codes, without the need for extensive manual annotation^18^.

One potential application for these methods is in clinical trial recruitment, which requires a comprehensive evaluation of both clinical trial eligibility criteria and patient medical histories in order to appropriately match patients who meet trial requirements^19–21^. A recent study developed an LLM framework that used GPT-4 to predict patient eligibility on a criterion-level basis with explanations and achieved near expert-level performance^22^. Recent work comparing proprietary and open-source models suggested that distillation along with fine-tuning can improve performance of open-source LLMs for patient trial matching, approaching that of GPT-4^23^. As opposed to Nievas et al.^23^, we used an open source model to generate the synthetic data, generated our data with MIMIC-III notes, and fine-tuned with QLoRA^24^. The fine-tuned models were evaluated against both the data used to create the synthetic question-answer pairs (MIMIC-III) as well as external data. Additionally, it is critical to use open-source models, even as the teacher. Deploying a model fine-tuned on GPT-4 outputs is likely against OpenAI’s terms of service^25^ as this would be deemed competing with OpenAI. As a whole, these developments show promise for the capacity of LLMs to aid in clinical information extraction for patient-trial matching but we observe in multiple evaluations performance is substantially higher when asking the model to answer single-order questions (e.g., what was the patient’s highest creatinine value?) as opposed to questions which require multiple steps (e.g., does this patient fit this trial’s eligibility criteria?).

In this work we demonstrate the ability to perform synthetic data distillation for scalable clinical note annotation, using a large open-source model to generate realistic questions based on patient clinical records which can be used to train a smaller model that can perform inference. Additionally, we perform an ablation study to understand which types of synthetic data yield optimal performance and we conduct comprehensive evaluations against multiple datasets. This is critical, because we observe it is substantially easier to achieve strong performance against synthetic data with manual review as opposed to fully human generated evaluations. Alongside the work, we release source code which provides a framework for meaningful, clinical information extraction synthetic data generation (https://github.com/bbj-lab/clinical-synthetic-data-distil) and an annotation tool built around making the annotation process faster particularly when LLM predicted annotations are already available (https://github.com/bbj-lab/annotation-ui). We are also releasing two newly manually annotated datasets to Physionet, which will be available via the same data use agreement as MIMIC-III/IV : 1.) **Annotated Synthetic Trial Criteria Questions**: 1,000 questions generated by the large 70B model as Synthetic Data, which have been human-reviewed, and 2.) **Apixaban Trial Criteria Questions:** 2,300 questions based on trial criteria from the ARISTOTLE apixaban clinical trial^26,27^.

## Results

The process of knowledge distillation by generating synthetic question and answer pairs using a large model (Llama 3.1 70B-Instruct) to teach a smaller model (Llama 3.1 8B-Instruct) is described in Figure 1. This process worked by passing in a discharge summary to Llama 3.1 70B-Instruct along with prompt instructions (Available in Supplementary Table 1 and within source code) to create questions meeting specific criteria (e.g., yes/no, numeric, or questions that cannot not be answered based on the content of the note). In addition to questions, the model was tasked with providing the section of the discharge summary an answer could be found (e.g., Pertinent Results), the source or exact text that allowed the model to answer the question, as well as an explanation of why the answer was correct based on the source and rest of the note. The model was also tasked with estimating the difficulty of the question it created (Supplementary Table 2).

**Figure 1.**
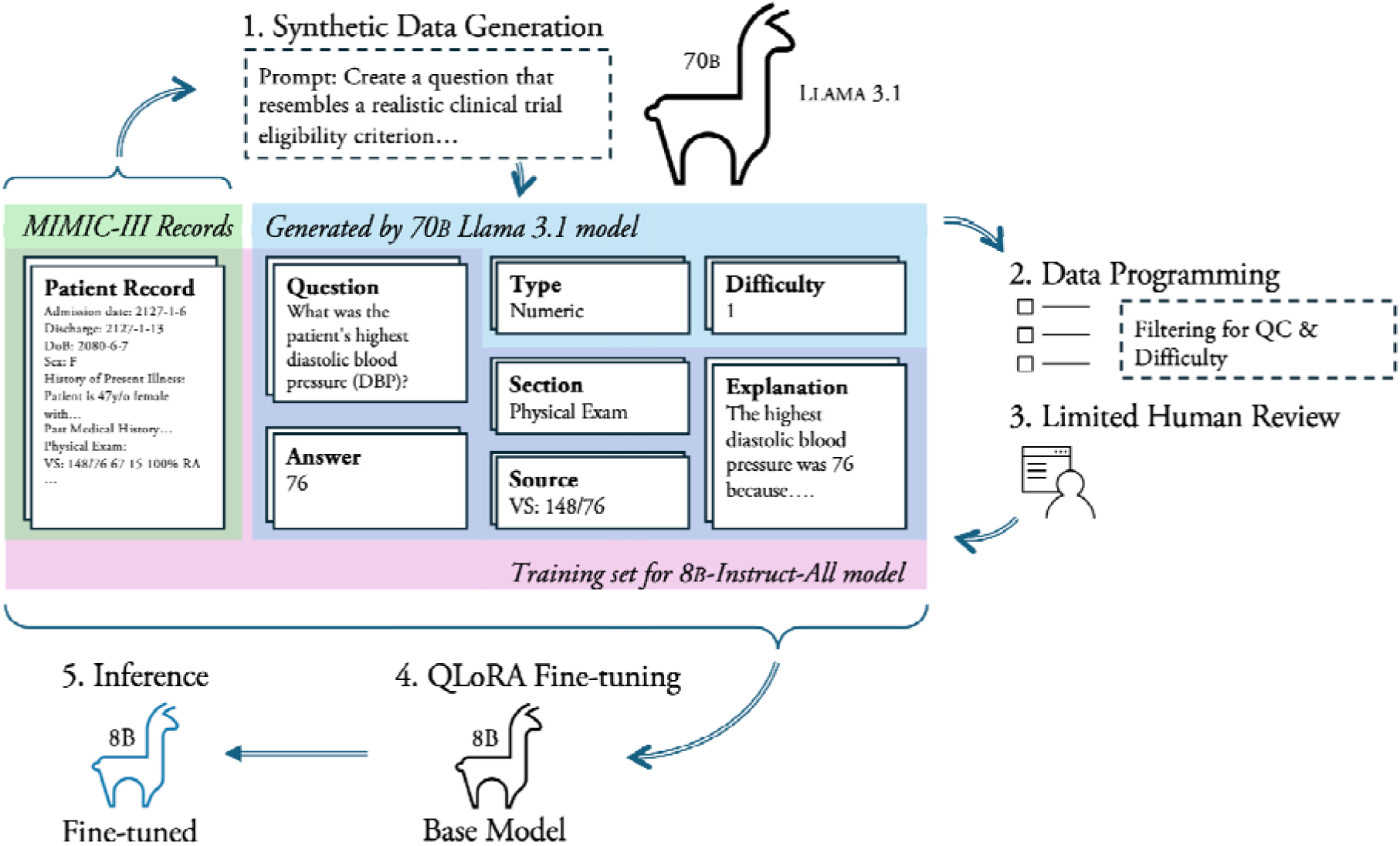
Synthetic Distillation Training Workflow. MIMIC-III records, outlined in green, are provided to the 70B-parameter Llama-3.1 model, which in turn generates the elements outlined in blue. After post-processing, the elements outlined in purple are provided to the 8B-parameter Llama-3.1 model for fine-tuning.

Next, these questions were filtered depending on which model was being fine-tuned (Table 1). For example, 8B-All includes all of the generated synthetic question and answer pairs, 8B-H-25K includes only the 25,000 questions the 70B-Instruct model ranked hardest within each category, 8B-NB-Only includes the 25,000 hardest numeric and boolean (yes/no) questions, and 8B-No-S includes the 25,000 hardest questions of each type but does not finetune on any of the supporting information, namely the explanation, the section the model believed the answer was in when generating the question, or the exact text which allowed for the model to answer the question (source). Next, QLoRA fine-tuning (detailed in Methods) was performed for each of the question categories to result in four fine-tuned models (8B-All, 8B-H-25k, 8B-NB-Only, and 8-No-S) in addition to the 2 instruct models open-sourced by Meta (8B-Instruct and 70B-Instruct) (Table 1).

**Table 1.**
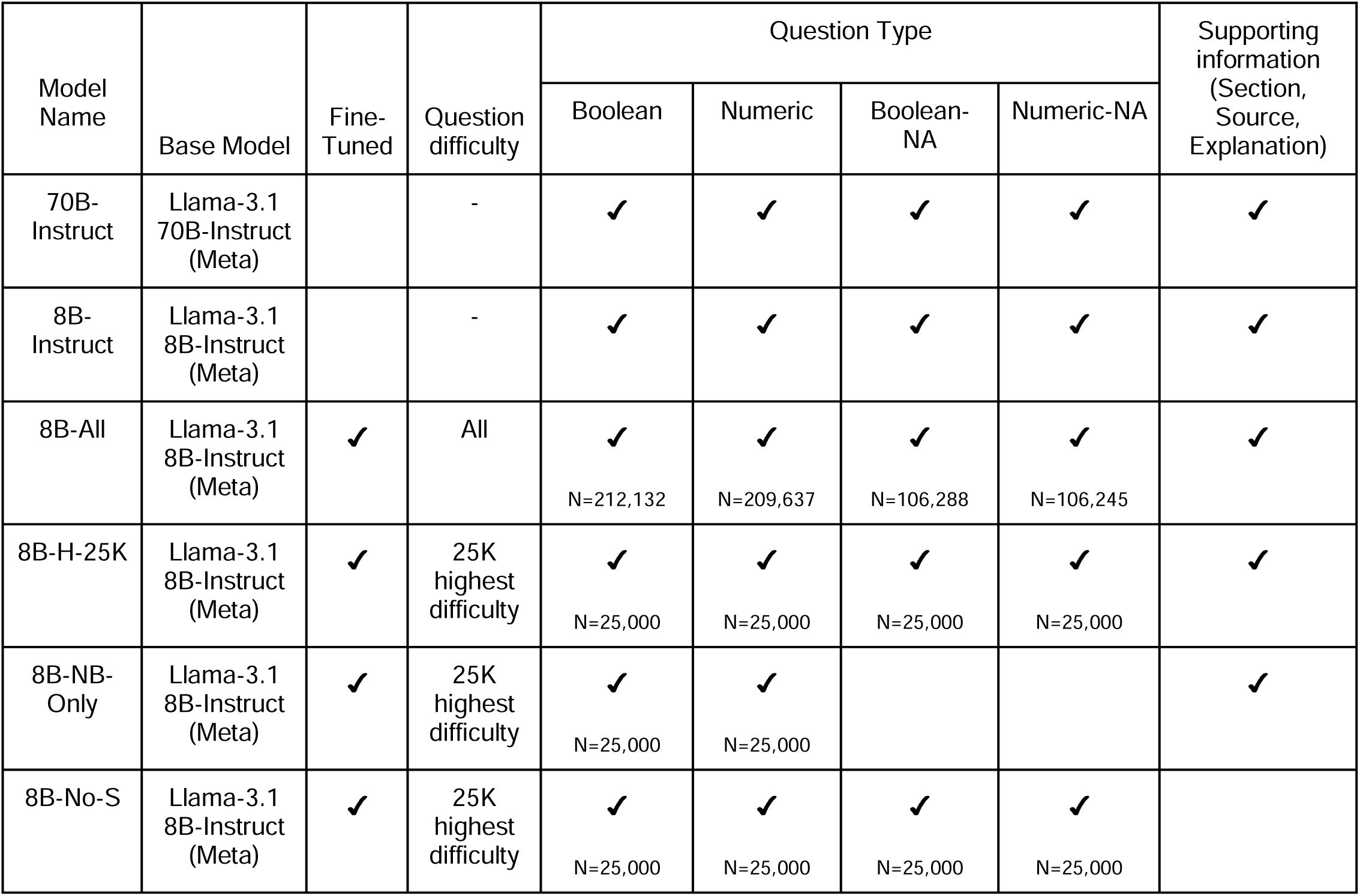
Comparison of the different models which were compared throughout the clinical information extraction tasks.

Each model was evaluated on three tasks: (i) annotated synthetic trial criteria questions, (ii) i2b2 Clinical Trial Eligibility Criteria Cohort Selection shared task from the 2018 National NLP Clinical Challenges, and (iii) apixaban trial criteria. We report performance metrics including Balanced Accuracy, which measures the average between sensitivity and specificity and can be used on imbalanced datasets, and Micro-F1 score. Micro-F1 was the primary metric used to judge the challenge, which permits direct comparison between our results and challenge entries (for the test set).

### Synthetic Data Evaluation

We evaluated the performance of the 8B-Instruct, 70B-Instruct, and the fine-tuned models on a manually annotated subset of 1,000 generated examples from the hold-out test set described in the methods datasets subsection (Table 2). The 8B-All model achieves the best overall accuracy (89.30%), outperforming even the 70B-Instruct model used for creating the synthetic data (76.20%). This was especially visible in the “NA” categories, where there appears to be a strong impact of training models explicitly on questions that cannot be answered based on the context (note) provided. Within each category, 8B-All and 8B-H-25k improved over 8B- Instruct, reflecting the impact of fine-tuning. 8B-H-25k also outperformed 70B-Instruct overall, suggesting that while the model benefits from further fine-tuning, a relatively small dataset of 25k examples can still provide an appreciable benefit. Unsurprisingly, the 8B-NB-Only model which was not fine-tuned on any “NA” data struggles in both of the NA columns, but it does perform very well for numeric and boolean and is actually the top performer for numeric questions.

**Table 2.** Model Accuracy on a subset of manually annotated Synthetic Labels (70B). Reported values include the mean accuracy and 95% CI.

|  | Accuracy Reported by Question Type |  |  |  |  |
| --- | --- | --- | --- | --- | --- |
|  | NA - Boolean<br>(N = 241) | NA - Numeric<br>(N = 232) | Numeric<br>(N = 236) | Boolean<br>(N = 291) | All Questions<br>(N = 1000) |
| 70B-Instruct | 69.5%<br>(63.5%, 75.1%) | 81.8%<br>(76.7%, 86.6%) | 61.7%<br>(55.5%, 67.8%) | **88.6%**<br>(84.9%, 92.1%) | 76.1%<br>(65.6%, 85.6%) |
| 8B-Instruct | 27.7%<br>(21.6%, 33.6%) | 78.4%<br>(73.3%, 83.2%) | 79.2%<br>(74.2%, 84.3%) | 87.3%<br>(83.5%, 91.1%) | 68.4%<br>(42.9%, 85.0%) |
| 8B-All | **88.0%**<br>(83.8%, 91.7%) | **98.3%**<br>(96.6%, 99.6%) | 83.9%<br>(78.8%, 88.1%) | 84.7%<br>(80.8%, 88.7%) | **89.1%**<br>(84.3%, 95.4%) |
| 8B-H-25k | 80.4%<br>(74.9%, 86.1%) | 85.5%<br>(80.6%, 90.1%) | 84.2%<br>(79.2%, 88.6%) | 88.0%<br>(84.2%, 91.1%) | 84.60%<br>(79.9%, 90.3%) |
| 8B-No-S | 78.9%<br>(73.8%, 83.8%) | 89.3%<br>(85.3%, 93.1%) | 80.6%<br>(75.4%, 85.2%) | 83.5%<br>(79.4%, 88.0%) | 83.0%<br>(79.7%, 87.1%) |
| 8B-NB-Only | 0.0%<br>(0.0%, 0.0%) | 40.0%<br>(33.6%, 46.6%) | **84.4%**<br>(79.2%, 88.6%) | 87.6%<br>(83.5%, 91.1%) | 54.0%<br>(22.2%, 87.0%) |

### i2b2 Clinical Trial Eligibility Challenge Evaluation

We next evaluated the performance of all base and fine-tuned models on the i2b2 2018 Clinical Trial Eligibility Challenge (Figure 2). Because we did not train on or otherwise use these data in our fine-tuning process we were able to assess the performance of models across both i2b2 train and test data.

**Figure 2.**
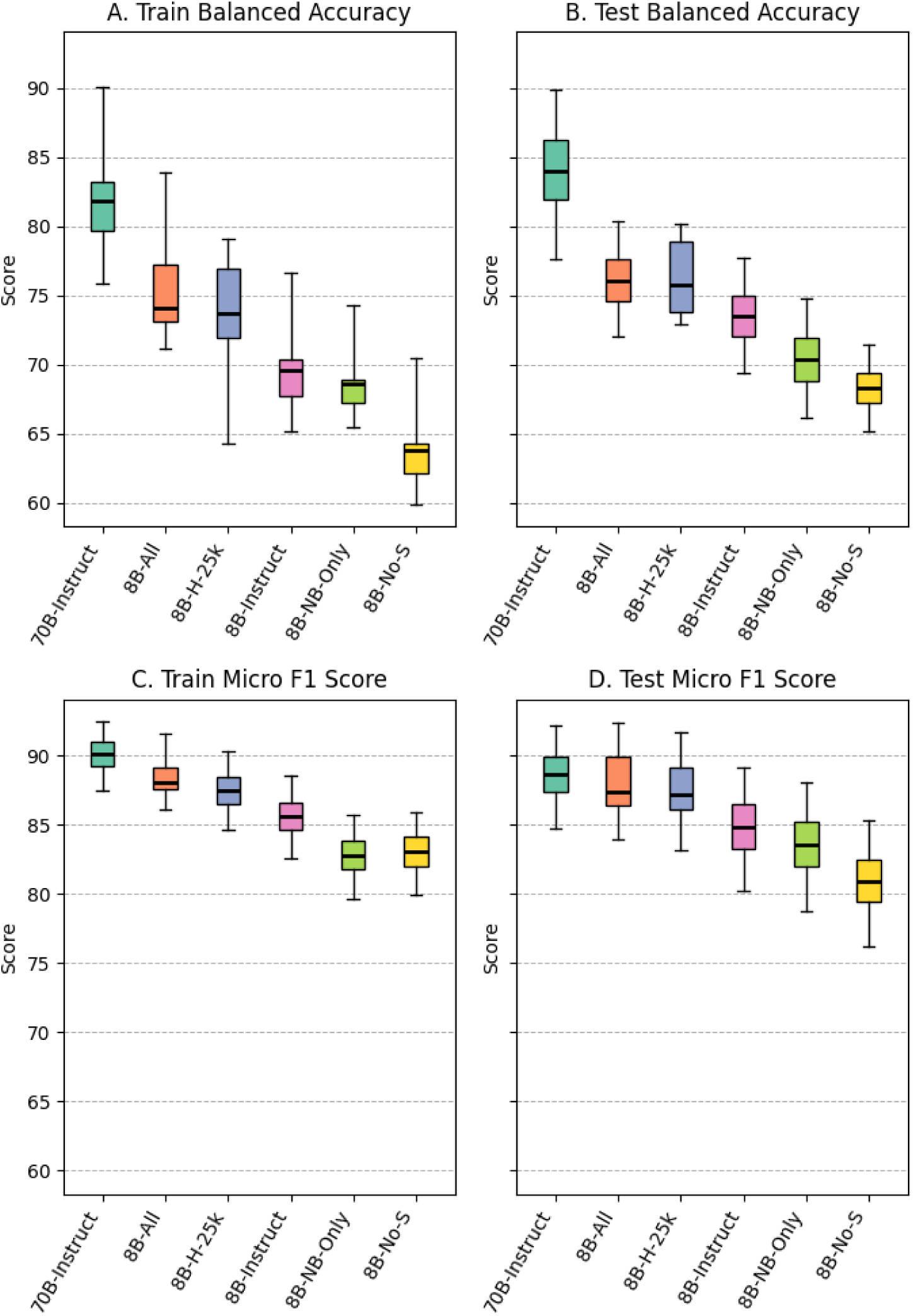
Comparison of model performance for the i2b2 (n2c2) Clinical Trial Eligibility Challenge. Evaluation includes the Training Set (A & C) because these data were not included during any of the pre-processing, hyperparameter selection or fine-tuning process of the models. All evaluations are zero-shot, but performance on Training (A & C) are separated from Test set (B & D) for clarity.

We evaluated two different values of two parameters, temperature and top_p (see Methods). We had a hypothesis that sampling strategies (i.e., higher temperature) might work well to force the model to provide an answer that aligned well with the explanation. However, we observed that the temperature did not not have a big impact, and a temperature of 0 slightly outperformed a higher temperature (Supplementary Table 3). The 70B-Instruct model performed the best on both train and test data. The two fine-tuned models which included all types and supporting information (8B-All and 8B-H-25K) outperformed the base 8B-Instruct model. The fine-tuned models that either did not include all types (8B-NB-Only) or did not include supporting information (8B- No-S) had worse performance than the base 8B-Instruct model.

An interesting trend we observed throughout this work was the need to isolate criteria and thus the prompts provided to the models into questions which required only single order answers. This was illustrated when comparing the performance of both the base models and fine-tuned models for their ability to either a.) directly answer a prompt question for a given criterion (i.e. direct boolean “yes” or “no”) vs. b.) extracting the numeric value relevant to the criterion and then performing post-processing to arrive at a boolean “yes” or “no” answer (Table 3). Within the i2b2 n2c2 challenge, two questions asked whether labs were abnormal (serum creatinine and hemoglobin levels). Across all models, numeric extraction followed by post-processing achieved higher performance compared to asking the model to directly answer the question.

**Table 3.** Comparison between directly answering clinical trial criteria about laboratory value ranges vs. extracting a number and applying rules-based post processing to determine whether to answer “yes” or “no” (i.e., ask the model to return a number, if that number is above a range answer yes, otherwise answer no).

| Criterion Title | Prompt Type | Prompt Question | Extracted Value Processing | Performance |  |  |  |  |  |
| --- | --- | --- | --- | --- | --- | --- | --- | --- | --- |
|  |  |  |  | Balanced Accuracy |  |  | Micro-F1 |  |  |
|  |  |  |  | 70B | 8B | 8B-All | 70B | 8B | 8B-All |
| Creatinine | Numeric | What was the patient's highest recorded creatinine level? Answer NA if there are no values. | <= 1.3: No<br>> 1.3: Yes<br>(Does not account for Sex) | 0.893 | 0.870 | 0.894 | 0.878 | 0.844 | 0.899 |
|  | Boolean | Has the patient ever had a serum creatinine level above the upper normal limit? (Typically > 1.3 mg/dL for men and 1.1 mg/dL for women). | None | 0.825 | 0.763 | 0.819 | 0.788 | 0.715 | 0.791 |
| HbA1c | Numeric | What was the patient's highest recorded hemoglobin A1c (HbA1c) value? Answer NA if there are no values. | >= 6.5: Yes<br>Else: No | 0.949 | 0.783 | 0.896 | 0.937 | 0.729 | 0.875 |
|  | Boolean | Has the patient ever had a hemoglobin A1c (HbA1c) level between 6.5 and 9.5 inclusive? | None | 0.774 | 0.583 | 0.743 | 0.774 | 0.462 | 0.760 |

### Apixaban Trial Criteria Evaluation

As the third evaluation task, we compared the performance of the base and fine-tuned models using manual annotations based on 23 questions resembling eligibility criteria from the apixaban clinical trial for a random sample of 100 patient notes from MIMIC-IV (Table 4). The fine-tuned 8B-All model achieved high performance exceeding Balanced Accuracy and Micro-F1 of 0.8 across all criteria assessed, with an overall average Balanced Accuracy of 0.93 and Micro-F1 of 0.94. This fine-tuned model outperformed the 8B-Instruct (Balanced Accuracy = 0.84, Micro-F1 = 0.86) and even the 70B-Instruct model (Balanced Accuracy = 0.89, Micro-F1 = 0.92). The model fine-tuned on the most difficult 25,000 questions, 8B-Instruct-H-25K, achieved a similarly high performance across criteria (average Balanced Accuracy = 0.95, Micro-F= 0.94), suggesting that either less total questions may be needed for fine-tuning, or that more difficult questions offer greater value in fine-tuning.

**Table 4.** Performance on clinical trial eligibility criteria for Apixaban.

|  | Balanced Accuracy |  |  |  | Micro-F1 |  |  |  |
| --- | --- | --- | --- | --- | --- | --- | --- | --- |
| Criterion | 8B-Instruct | 70B-Instruct | 8B-All | 8B-H-25k | 8B-Instruct | 70B-Instruct | 8B-All | 8B-H-25k |
| AST | 54.0% | 97.0% | 94.3% | 99.4% | 0.54 | 0.97 | 0.94 | 0.99 |
| Bilirubin | 100.0% | 99.0% | 100.0% | 99.3% | 1 | 0.99 | 1 | 0.99 |
| Creatinine | 85.0% | 80.0% | 84.0% | 85.0% | 0.85 | 0.8 | 0.84 | 0.85 |
| Hemoglobin | 96.0% | 90.0% | 98.0% | 96.0% | 0.96 | 0.9 | 0.98 | 0.96 |
| Platelets | 78.0% | 87.0% | 79.6% | 79.1% | 0.75 | 0.87 | 0.8 | 0.81 |
| AFib | 98.0% | 81.7% | 98.0% | 98.0% | 0.97 | 0.84 | 0.97 | 0.97 |
| Ablation for Afib | 89.5% | 64.6% | 81.8% | 98.0% | 0.98 | 0.94 | 0.96 | 0.96 |
| Arterial Hypertension | 99.4% | 95.0% | 97.7% | 97.1% | 0.99 | 0.98 | 0.96 | 0.95 |
| Bipolar Disorder | 100.0% | 100.0% | 100.0% | 100.0% | 1 | 1 | 1 | 1 |
| Bleeding | 91.6% | 92.1% | 89.4% | 80.0% | 0.91 | 0.85 | 0.89 | 0.8 |
| Blood Glucose | 25.0% | 84.0% | 98.5% | 94.0% | 0.25 | 0.84 | 0.98 | 0.94 |
| Chads2 | 89.0% | 94.0% | 94.9% | 97.6% | 0.89 | 0.94 | 0.95 | 0.98 |
| Heart Failure | 96.5% | 98.0% | 98.2% | 99.0% | 0.96 | 0.98 | 0.98 | 0.99 |
| Hemorrhagic Tendencies | 60.3% | 62.7% | 85.6% | 96.1% | 0.52 | 0.78 | 0.89 | 0.92 |
| Left ventricular ejection fraction | 72.0% | 90.0% | 94.9% | 96.0% | 0.72 | 0.9 | 0.95 | 0.96 |
| Depression | 100.0% | 95.3% | 98.7% | 97.5% | 1 | 0.92 | 0.98 | 0.96 |
| Makes Medical decisions | 79.5% | 81.4% | 95.3% | 93.4% | 0.91 | 0.9 | 0.91 | 0.93 |
| Peptic Ulcer Disease | 75.0% | 99.5% | 92.9% | 99.5% | 0.94 | 0.99 | 0.99 | 0.99 |
| Prior Stroke | 81.7% | 86.2% | 88.5% | 89.2% | 0.89 | 0.92 | 0.94 | 0.94 |
| Recent Stroke | 75.3% | 94.2% | 94.2% | 94.2% | 0.86 | 0.89 | 0.89 | 0.89 |
| Schizophrenia | 100.0% | 100.0% | 100.0% | 100.0% | 1 | 1 | 1 | 1 |
| Valvular Disease | 87.2% | 83.9% | 86.8% | 86.8% | 0.95 | 0.92 | 0.93 | 0.93 |
| requiring Surgery |  |  |  |  |  |  |  |  |
| Diabetes | 98.3% | 82.2% | 99.1% | 98.3% | 0.97 | 1.00 | 0.99 | 0.98 |
| Average | 84.0%<br>(75.9%,<br>90.5%) | 88.7%<br>(84.3%,<br>92.6%) | 93.5%<br>(90.9%,<br>96.1%) | 94.2%<br>(90.9%,<br>96.9%) | 0.863<br>(0.783-0.934) | 0.904<br>(0.873-0.934) | 0.945<br>(0.920,0.96<br>6) | 0.943<br>(0.912,<br>0.971) |

There were some criteria where base 8B-Instruct model had relatively lower performance, including extraction of aspartate aminotransferase (AST) (Balanced Accuracy = 0.54, Micro-F1 = 0.54), blood glucose (Balanced Accuracy = 0.25, Micro-F1 = 0.25), and left ventricular ejection fraction (Balanced Accuracy = 0.72, Micro-F1 = 0.72). The use of the larger 70B-Instruct model dramatically improved performance for these criteria, exceeding Balanced Accuracy and Micro-F1 of 0.84. The fine tuned models 8B-All and 8B-H-25k performed comparably to the 70B model, and in some cases outperformed it. All three models for the AST criteria led to Balanced Accuracy and Micro-F1 scores of 0.94 and above. For blood glucose, the fine-tuned models 8B-All (Balanced Accuracy = 0.98, Micro-F1 = 0.98) and 8B-H-25k (Balanced Accuracy = 0.94, Micro-F1 = 0.94) achieved higher performance than the 70B-Instruct model (Balanced Accuracy = 0.84, Micro-F1 = 0.84). For identification of hemorrhagic tendencies, the model fine-tuned on the 25k most difficult questions led to the biggest performance improvement (Balanced Accuracy = 0.96, Micro-F1 = 0.92) compared to both the 8B-All (Balanced Accuracy = 0.96, Micro-F1 = 0.92) and 70B-Instruct models (Balanced Accuracy = 0.96, Micro-F1 = 0.92).

For some criteria, the 70B-Instruct model did not perform as well as any of the 8B-Instruct models, including the base model. This was the case when detecting the presence of atrial fibrillation (**8B-Instruct:** Balanced Accuracy = 0.98, Micro-F1 = 0.97; **70B-Instruct:** Balanced Accuracy = 0.65, Micro-F1 = 0.84) and whether there was planned/past ablation for atrial fibrillation (**8B-Instruct:** Balanced Accuracy = 0.89, Micro-F1 = 0.98; **70B-Instruct:** Balanced Accuracy = 0.65, Micro-F1 = 0.94). There were also some criteria, including creatinine and platelets, where the models did not perform as well as other criteria as no model exceeded 0.85 for either balanced accuracy or micro-F1. Of the manually annotated notes, 60% did not have a numeric value for platelet count available in the note, while only 3% did not have a serum creatinine value available (Supplementary Table 4). This rate may be at least in part due to the fact that the de-identification process for MIMIC-III seemed to accidentally redact some platelet values. During the manual annotation process we did not observe this occurring with other laboratory values.

### Resource requirements

Data distillation allowed the models to be run with vastly reduced resource requirements compared to the 70B- Instruct model. All model evaluation was done on the Center for Research Informatics’ “Randi” cluster at the University of Chicago. The cluster’s GPU nodes each contain 8 Nvidia A100 GPU’s with two 16-core 3.0-GHz AMD Milan processors. We monitored seconds/example, tokens in/second, and tokens out/second for both the 8B-parameter and 70-B parameter architectures and reported these in Figure 3. These differences could translate into meaningful cost savings. For example, performing a study of the Apixaban criteria (23 questions) for 10,000 patients to identify a cohort on the least expensive cloud provider would be $3,132 less expensive for the 8B vs. 70B parameter models (see Supplementary Table 5 for a comparison of current rates among the main providers). In this example, running the 8B-parameter model would cost less than $1,000 (0.535 sec./ex. * 230k ex. * 1/3600 hr./sec. * $27.2/hr. = $929), while the 70B-parameter model would cost over $4,000 (2.34 sec./ex. * 230k ex. * 1/3600 hr./sec. * $27.2/hr. = $4066).

**Figure 3.**
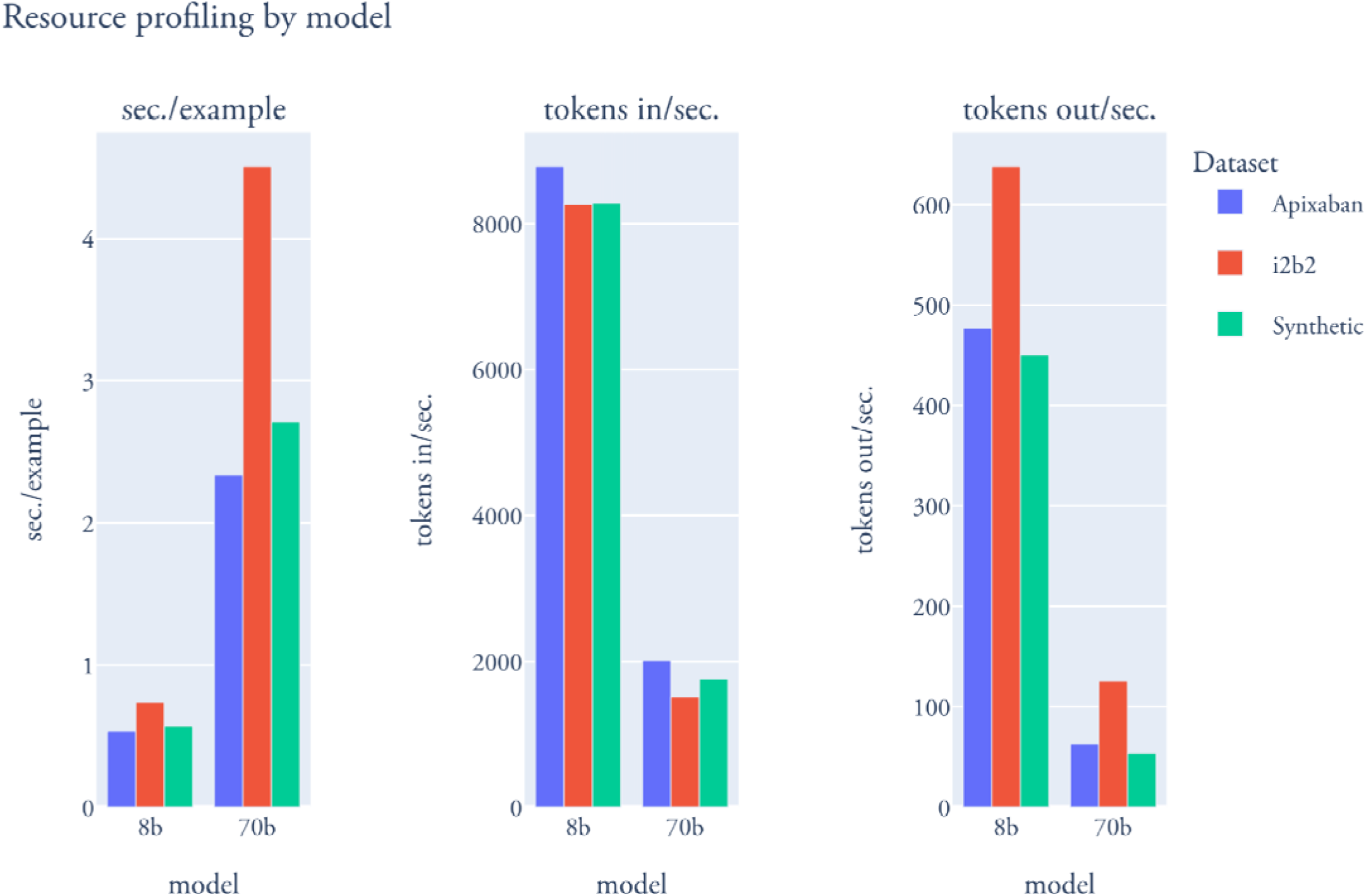
Comparison of inference speed across model sizes and evaluation tasks.

## Discussion

In this study, we present an approach to improve the scalability of open-source LLMs for clinical information extraction using synthetic data distillation. We used the larger Llama-3.1-70B-Instruct to generate synthetic data, consisting of question-answer pairs with supporting information and difficulty scores. These were used to fine-tune the smaller Llama-3.1-8B-Instruct model. We also explored the impact of fine-tuning on different amounts and subsets of synthetic data (including one fine-tuned with all data, one fine-tuned with only the hardest 25K questions, one fine-tuned without questions where the note does not contain the answer - NA, and one fine-tuned without any supporting information). We observe that the inclusion of NA and supporting information was critical to the high performance of fine-tuned models especially when applied to fully human generated evaluations as opposed to synthetic data with human review. When evaluating the accuracy of these models based on manually annotated synthetic data, we found that the model fine-tuned on all synthetic data (8B-All) achieved a high overall accuracy that exceeded that of a larger base model (70B-Instruct). We found that these fine-tuned models also performed well across different clinical tasks, including the i2b2 Clinical Trial Eligibility Challenge and a dataset designed to resemble real eligibility criteria from the apixaban clinical trial. The fine-tuned models can achieve performance comparable to, and in some cases exceeding, that of even the larger model that served as the teacher. Even when fine-tuning is performed using only a subset of the hardest questions in the synthetic dataset, the performance still improves over base models, suggesting that targeted fine-tuning with less data can still be beneficial. Finally, we release several artifacts we believe will be beneficial to researchers further developing approaches for clinical information extraction: a.) source code - both the framework for synthetic data generation for clinical information extraction model fine-tuning as well as the annotation tool which allowed for faster manual review of LLM pre-annotated notes, and b.) datasets - two manually annotated datasets (Annotated Synthetic Trial Criteria Questions and Apixaban Trial Criteria Questions) which will allow for researchers to evaluate future methods for clinical information extraction.

The use of LLMs to extract information from clinical notes has already demonstrated the potential to improve upon traditional methods relying on rule-based methods or extensive manual annotation. While proprietary models, such as GPT-3 and GPT-4, have shown strong performance for this purpose, their deployment in healthcare settings can be limited by computational costs and licensing barriers^22^. Our findings align with recent research suggesting that fine-tuning open-source models with synthetic data can improve their performance across clinical information extraction tasks, bringing it closer to that of proprietary models^23^. By generating synthetic data, this approach also reduces reliance on manually labeled data. The use of smaller, open-source models that can perform well opens the door for broader adoption of LLMs in the healthcare settings in a cost-effective and privacy-compliant manner. The reduced computational requirements of these smaller models can make them more accessible to hospitals with limited healthcare IT infrastructure. Another advantage of this approach is the adaptability, as smaller models can be better tailored to the specific needs of individual hospitals. As more resource-efficient LLMs such as Llama 3.2 continue to evolve, we can extend this approach to even smaller models^28^.

By enabling scalable information extraction from unstructured notes, this approach presents a promising opportunity for retrospective research through its potential impact on enhancing patient phenotyping. This is particularly important when studying complex and heterogeneous patient populations, where phenotyping approaches relying solely on structured data, such as ICD codes, fall short. Better phenotyping can result in improved quality and relevance of retrospective studies.

While this has exciting potential, we also note some of the limitations and challenges identified through manual review of the synthetic data generated by Llama-3.1-70B-Instruct that may begin to elucidate failure modes for these models. In general, the model struggled with ranges when forming numeric questions. In multiple instances, a range (e.g. 60-70%) would be collapsed to one of its limits (60% or 70%) in a numerical answer. In at least one instance, the model had difficulty comparing a range and a given value outside that range (e.g. concluding that >70% precludes 50%). This contrasts with the model’s generally consistent ability to locate the highest or lowest value in a sequence of measurements (e.g. finding the highest blood pressure recorded in a note containing multiple readings).

The model also sometimes struggled with redacted data and contextual understanding. In one instance, the model identified numbers in a redaction tag as the answer to a question. This tag would have contained the correct answer prior to redaction. The tag itself, “[**3-22**]”, contained numbers and this may have contributed to the model’s confusion. In another example, the model successfully identified the inappropriately partially- redacted “[churgg [**Doctor Last Name **] disease” as Churg-Strauss disease. In another case, the model correctly identified a patient’s hemoglobin value but then incorrectly concluded that it fell below the normal range. This conclusion would have been correct had the patient been male; however, the patient was female and the reference range is lower for females. In another case, the model asked if a female over 70 was “a candidate for future pregnancy?” Interestingly, the model was also able to identify and parse a fishbone diagram within a note correctly answering questions about lab values contained within the diagram.

The model also sometimes lacked creativity when generating questions with unspecified answers. To generate questions that could not be answered using the contents of a note, the model seemed to commonly inquire about BMI (height and weight measurements are recorded separately from these notes and so are often not contained in the text) and the results from a 6-minute walking test (6MWT). In the full test set containing 42,498 instances, we found 997 questions related to BMI (98.7% of which resolved n/a) and 666 questions related to a 6-minute walk test (all of which resolved n/a). The model would also ask about measurements from a patient prior to them seeking medical attention, which are typically unavailable in these notes. Additionally, the models would sometimes struggle with repetitive generation. In the test set, we found 1,676 (3.94%) questions containing “creatinine”. Admittedly, our prompt for numeric type questions included an example “What was the patient’s highest creatinine measurement recorded in the note?” However, a majority (1,151) of these questions were of na-numeric, boolean, or na-boolean type, and none of those prompts mention creatinine.

We found that carefully worded prompts could help to avoid some of the issues described in the previous section. By rewording questions, we could deter the model from drawing inferences and obtain less ambiguous question-answer pairs. For questions that asked if a patient had a history of X, where X was not mentioned in the note, the model would sometimes conclude that a patient did not have a history of X because X was not mentioned in the note, and other times conclude that the question could not be definitively answered from the contents of the note. This ambiguity could be resolved by modifying the question to ask if a patient’s history of X could be found in the note. This is especially critical because it allows us to use a combination of clinical expertise and post-processing to knowingly make assumptions where appropriate about whether X would have been in the note if they had it, as opposed to the model making this assumption for us without our knowledge. In a similar vein, questions on the highest recorded value of Y could be reworded to ask about the highest value of Y found in the note. Instead of asking if a patient’s measurement for Z fell within a normal range, we could ask the model to return the patient’s measurement for Z and then evaluate if Z fell within the standard reference range as a separate step. This allowed us to avoid having the model reason about ranges of values, a known area of difficulty.

Developing resource-efficient LLMs to extract relevant information from clinical notes is a rapidly advancing discipline with many open questions. For example, there may be better ways to make the distillation process more data-efficient. In this work, we showed how fine-tuning on only a fraction of the synthetic dataset (e.g. 8B-H-25k) still appreciably enhances the base 8B-Instruct model. Different criteria for selecting a subset of the fine-tuning data may better maintain performance while decreasing data requirements^29,30^. Ordering the fine-tuning set by increasing difficulty and interleaving question types may also help^31^.

Future work could consider whether a metric besides micro-F1 could better characterize good performance. We used micro-F1 in part because it benchmarked the original i2b2 challenge. However, some researchers view patient-clinical trial matching as a ranking problem and consequently report metrics like normalized discounted cumulative gain at k and precision at k^22,23^. We could also consider the optimal way to handle ambiguity in notes. Unlike tabular or structured data that typically complies to a strict format, notes often include estimates and conjectures, especially when discussing medical history. There are often question marks next to past diagnoses and values reported. Another potentially interesting extension of this work can look into how data from multiple notes could be combined. Many people have a medical history spanning decades. For selection criteria involving disease progression or patient history, multiple notes may be required to obtain a complete answer. Combining records in a time-aware manner remains an open problem.

Synthetic data distillation and fine-tuning of smaller, open-source LLMs that can be locally deployed within existing healthcare IT infrastructures can serve as a scalable alternative to more resource-intensive, proprietary models for clinical information extraction. The ability for scalable extraction of information from unstructured clinical notes allows for broader adoption in diverse healthcare system settings, with the potential to strengthen retrospective research by enabling more precise and accurate phenotyping. This work contributes to efforts to support the effective and practical integration of LLMs in healthcare settings, with the ultimate goal of supporting medical research to improve patient outcomes.

## Methods

### Base synthetic data distillation

In this section, we describe our knowledge distillation process which uses a large model, Llama-3.1-70B- Instruct, to generate training examples for the smaller model, Llama-3.1-8B-Instruct (Figure 1):

#### 1. Synthetic Data Generation

For each patient record, we used Llama-3.1-70B-Instruct^32^ to generate different, patient note-specific questions similar to clinical trial eligibility criteria of a given type (Exact language available in Supplementary Table 1 and source code). We prompted the model to supply its answers in json format. Each json includes the following: (1) the *question*; (2) the *question type*; (3) the *answer*, (4) the *section* of the note containing the answer (*e.g.,* Past Medical History, Plan, etc.); (5) the verbatim *source* of the answer from the clinical note; (6) a *difficulty level* for the question on a scale of 1-10; and (7) an *explanation* justifying the answer choice, including how the source helped to answer the question.

We included the following question types: “boolean” (answer “Yes” or “No”), “numeric”, “na-boolean”, and “na- numeric”, where the “na” types corresponded to questions that could not be answered relying on the information in the note but seemed like they would be applicable to this patient and are similar to clinical trial eligibility criteria. For “na” type questions, we stipulated the section to be “Not Found” and the source was “Not in Note.” The purpose of the “na” types as well as the supporting data was to try to teach the model not to provide seemingly confident answers (i.e., hallucinations) when there doesn’t exist sufficient evidence in the note to draw a conclusion. See Supplementary Table 6 for example questions of each type. For the specific language used to generate each question type, including the specific example supplied in the prompt, see Supplementary Table 1.

We generated 212,132 boolean question and answer (Q&A) pairings, 209,637 numeric Q&A pairings, 106,288 “na-boolean” Q&A pairings, and 106,245 “na-numeric” Q&A pairings. The number of questions arose from running the synthetic data generation process on 10,000 discharge summaries, where the model was asked to generate 20 boolean questions (10 with yes as the answer and 10 with no), 20 numeric questions, and 10 of each “na” category. The model tended to provide slightly more than the requested number of questions per note. The number of questions of each type per difficulty score assigned by Llama-3.1-70B are described in Supplementary Table 2.

#### 2. Data Programming

For each question type, we select the 25,000 most difficult questions according to the LLM-estimated difficulty rating and randomly split them into a training and test set at a 90%-10% ratio. We perform post-processing to extract our requests from the json response and handle malformed json outputs. The datasets are randomly shuffled prior to fine-tuning.

#### 3. Limited Human Review

To ensure data quality for the fine-tuning process, we manually reviewed a random sample containing 1,000 questions generated by Llama-3.1-70B-Instruct. For this purpose, we developed an open-source tool that facilitates record review from within a web browser (https://github.com/bbj-lab/annotation-ui). Users with minimal technical experience can check patient records against the generated question-answer pairs and refine answers if needed. Statistics about the numbers of questions which required refinement are available in Supplementary Table 1. Manual review allowed us to both profile the accuracy of the synthetic data generation process and to better understand common failure modes.

#### 4. QLoRA Fine-tuning

After data programming and limited human review, we used the refined synthetic dataset to perform supervised fine tuning on an instance of Llama-3.1-8B^32^. Specifically, we fine-tuned with QLoRA^24^, a quantized version of Low-Rank Adaptation (LoRA:^33^). LoRA fine-tunes the attention weights in a pre-trained transformer with a low-rank update (a *d×k* matrix *BA*, where B is *d×r* and A is *r×k* where *r*≪*min{d,k}*) that significantly reduces the number of required parameters and does not add to inference latency. QLoRA operates on a quantized transformer, i.e. one that uses 4-bit as opposed to 16-bit parameters, to further reduce memory requirements and uses paged optimizers that manage the exchange of memory between GPU and CPU components.

#### 5. Inference - Sampling hyperparameter selection

During generation, we tested different values of *temperature* and *top_p* (specifically temperatures of 0 and 1 and top_p of 0.5 and 0.95). Temperature controls the randomness of sampling, with higher temperatures corresponding to more novelty in generated output. However, increasing temperature may also make text less coherent and hallucinations more likely. Consequently, higher values of temperature are often used for creative tasks, while lower values are used for dialoguing about matters of fact. Chang et al [2023]^34^ hypothesized that lower values of temperature may be better suited to question-answering with attribution. The top_p parameter controls nuclear sampling^35^, with higher values corresponding to a more permissive threshold for filtering.

Setting temperature = 0 and top_p = 1 results in a nearly deterministic, greedy sampling strategy that aims to select the most likely token given the current context. Setting temperature = 1 and top_p = 0.5 restricts tokens to come from a likely subset of the token set, but otherwise samples according to the predicted odds. We limit this parameter evaluation to the *i2b2 n2c2* challenge and report the full results for all parameters in Supplementary Table 3. Because we did not see a benefit when increasing the temperature we fixed temperature = 0 and top_p = 1 for all other evaluations.

### Versions of Fine-tuned Models (Ablation study on fine-tuning data selection - Table 1)

Each version fine-tuned the Llama-3.1-8B-Instruct released by Meta as the base model.

#### All (Labeled 8B-All)

Fine-tuning was performed with the complete dataset, using question, answer, question type, section, source, and explanation as described in the methods.

#### Hardest (Labeled 8B-H-25k)

To determine the performance impact of reducing training set size, we selected the 25,000 questions the model determined had the highest difficulty in Step #1 (*Synthetic Data Generation)* for each question type. This subset of the original training data was then used to fine tune the model.

#### Hardest Boolean and Numeric (Labeled 8B-NB-Only)

To determine the impact that n/a questions have on model fine tuning, we selected the most difficult 25,000 questions for only the boolean and numeric types (dropping “na-boolean” and “na-numeric”) from the original dataset for fine-tuning.

#### No Support (Labeled 8B-No-S)

To determine the usefulness of including textual references and an explanation of the correct answer, we dropped the section, source, and explanation from the original training set and fine-tuned the model with this data.

### Datasets

#### Annotated Synthetic Data

We evaluate methods on a held-out set of 42,498 synthetic examples generated in an identical manner to the dataset used for fine-tuning. The breakdown of examples by type was as follows: 10,722 (25.2%) boolean, 10,666 (25.1%) numeric, 10,664 (25.1%) na-boolean, and 10,446 (24.6%) na-numeric. From this set, we drew a random sample containing 1,000 examples and manually annotated it as described in the “Limited Human Review” subsection of our methods, correcting questions, answers, and explanations when necessary. We provided a summary of this dataset in Supplementary Table 1 and have released a copy of it on Physionet.

#### i2b2 2018 National NLP Clinical Challenges (n2c2): Cohort Selection

We evaluate methods on the clinical trial eligibility criteria cohort selection shared task from the 2018 National NLP Clinical Challenges^36^. Track 1 contains 288 de-identified longitudinal medical records for patients with diabetes, many of whom are at risk for heart disease. The records are manually annotated according to 13 selection criteria adapted from real clinical trials and split into a 202-patient training set and 86-patient test set. At the time of the challenge, the top-performing team adopted a rule-based method to obtain a micro-F_1_ score of 0.91 on the test set. Other teams achieved similar results (F_1_ > 0.9) with hybrid approaches; for example, cTakes ^37^ was used by 3 of the top 5 teams to extract knowledge from the text. Because we only use this dataset to test zero-shot extraction and do not train on it, we are able to evaluate the model performance on both the training and test sets to have a larger sample size.

#### Apixaban Clinical Trial

We also evaluate methods on clinical trial eligibility criteria resembling those of the 2011 ARISTOTLE trial comparing apixaban to warfarin^26^. We developed 23 human-generated boolean and numeric questions assessing these criteria (Supplementary Table 4). Using these questions, we manually annotated notes for 2300 total question-answer pairs within MIMIC-IV^38,39^. Notes from MIMIC-IV were taken from after 2012 to ensure no overlap with any of the notes from MIMIC-III which were used to generate synthetic data. We are releasing the dataset and manual annotations to Physionet and will make them available under the same data use terms as MIMIC-III/IV.

## Author Contributions

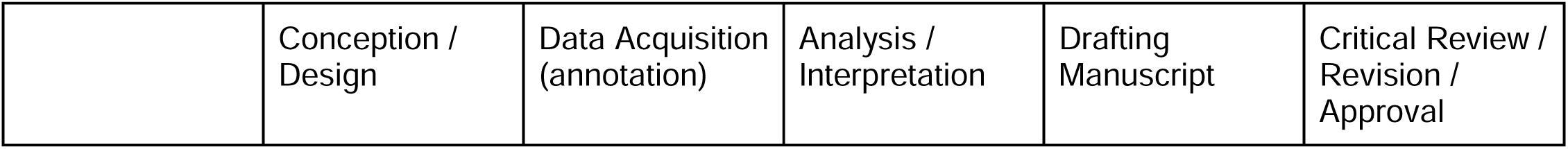
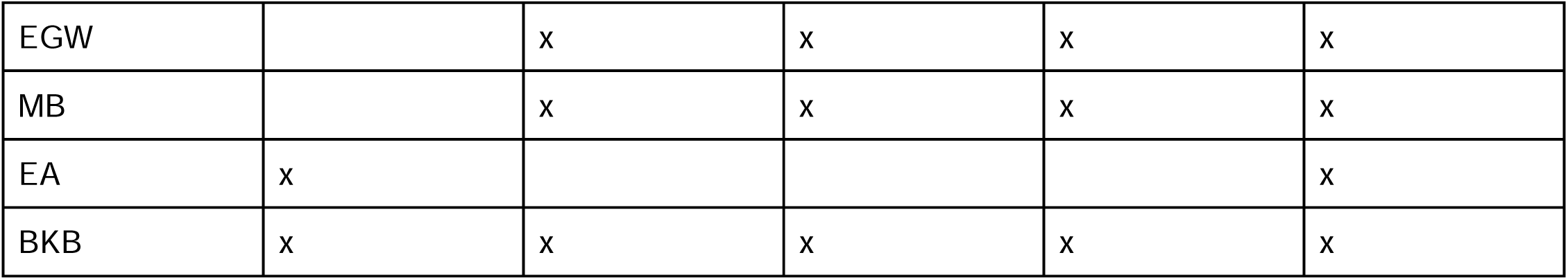

## Acknowledgements

This work was funded in part by the National Institutes of Health, specifically the National Institute of Neurological Disorders and Stroke grant number R00NS114850 to BKB. This project would not have been possible without the support of the Center for Research Informatics at the University of Chicago and particularly the High-Performance Computing team. The authors are grateful for the resources and support this team provided throughout the duration of the project. The Center for Research Informatics is funded by the Biological Sciences Division at the University of Chicago with additional funding provided by the Institute for Translational Medicine, CTSA grant number UL1 TR000430 from the National Institutes of Health.

## Data Availability

The MIMIC-III [Johnson, et al., 2016] and MIMIC-IV [Johnson, et al., 2023] datasets are available from PhysioNet. The Annotated Synthetic Questions and the Apixaban Trial Criteria Questions are in review with PhysioNet as described in the datasets subsection.

## Supplementary Information

**Supplementary Table 1.** Synthetic data format and annotation summary. We manually annotated 1000 fine-tuning examples generated by Llama-3.1-70B. Overall, 872 of the questions remained unchanged after manual review. A breakdown by question type occurs in the following table. The column “occurrence in data” corresponds to the frequency with which the question occurred in the dataset and the “percentage requiring editing” corresponds to how often that type of question needed to be edited. The “request provided” and “example provided” both correspond to sections in the prompt used during data generation.

| type | occurrence in data | percentage requiring editing | request provided | example provided |
| --- | --- | --- | --- | --- |
| boolean | 29.1% | 15.6% | Give a list of 10 total, different, patient note-specific questions similar to clinical trial eligibility criteria. 5 should have 'No' as the correct answer and 5 should have 'Yes' as the answer. | ````json [ { "question": "Does the note state that the patient is breathing normally on room air?", "type": "Yes/No", "answer": "No", "section": "History of Present Illness", "difficulty": "2", "source": "She currently is dependent on oxygen and wears 1.5-2 liters around the clock", "explanation": "The note states that she relies on oxygen and provides the amount as 1.5-2 liters so she is not breathing room air. We can assume since she is receiving o2 supplementation and dependent on it, she cannot breathe normally on room air." } ], ```` |
| numeric | 23.6% | 5.8% | Give a list of ten different, realistic, patient note-specific questions similar to clinical trial eligibility criteria with a numeric answer (only generate questions if numeric answers are appropriate, otherwise end the response). All questions should be specific, many numeric values can be listed more than once so make sure to specify first, last, at admission, on discharge, highest, lowest, on a specific date, within ED / ICU etc.. | ````json [ { "question": "What was the patient's highest creatinine measurement recorded in the note?", "type": "Numeric", "answer": "1.4", "section": "Pertinent Results", "difficulty": "4", "source": "12/03/2023: CREAT: 1.4 \n 12/07/2023: CREAT: 1.1", "explanation": "The highest CREAT measurement was 1.4 because the only other creatinine measurement was 1.1 on 12/07/2023." } ], ```` |
| na-boolean | 24.1% | 14.8% | Give a list of 5 Yes/No questions that are not answerable using the note. These should be questions which seem like they would be applicable to this patient and are similar to clinical trial eligibility criteria but cannot be answered based on the information in the note. These questions need to be things where the answer cannot be assumed simply because something is not mentioned (e.g., They should not be questions about whether the patient has been diagnosed with serious or chronic diseases because if they were it would be mentioned in the note, since it is not mentioned we can assume the answer is no rather than NA. Do not generate questions where Yes or No is known or can be inferred. | `'''json [ { "question" : "Does the note state the patient has ever taken aspirin for MI prevention?", "type": "Yes/No", "answer" : "N/A", "section": "Not Found", "source" : "Not in Note", "difficulty": "4", "explanation": "The note does not include medication history. It only includes medications prescribed during this encounter. If there were a medication history we would check the list to see if is present. If it was present we would answer Yes, if it were not present we would answer No but because there is no medication history we answer N/A" }, ]'''` |
| na-numeric | 23.2% | 13.4% | Give a list of 5 questions asking for numeric answers but where the note does not contain the answer. These should be questions which seem like they would be applicable to this patient and are similar to clinical trial eligibility criteria but cannot be answered based on the information in the note. | `'''json [ { "question" : "What was the patient's highest A1C recorded in the note during the hospitalization?", "type": "Yes/No", "answer" : "N/A", "section": "Not Found", "source" : "Not in Note", "difficulty": "4", "explanation": "The note does not include an A1C value during the hospitalization and we cannot infer a value for this patient." }, ]'''` |

**Supplementary Table 2.** Number of questions Llama 3.1 70B assigned each difficulty to during the synthetic data generation process.

| Difficulty | Boolean | Numeric | NA - Boolean | NA - Numeric |
| --- | --- | --- | --- | --- |
| 0 | 0 | 4 | 0 | 0 |
| 1 | 95,963 | 14,968 | 223 | 303 |
| 2 | 89,754 | 69,031 | 2,794 | 3,655 |
| 3 | 22,327 | 78,494 | 11,508 | 12,537 |
| 4 | 3,519 | 41,809 | 20,491 | 17,930 |
| 5 | 416 | 4,663 | 32,053 | 28,579 |
| 6 | 123 | 530 | 24,448 | 22,430 |
| 7 | 9 | 20 | 10,116 | 12,040 |
| 8 | 13 | 50 | 3,984 | 7,029 |
| 9 | 0 | 1 | 671 | 1,733 |
| 10 | 8 | 71 | 0 | 9 |

**Supplementary Table 3.** Performance on i2b2 2018 Clinical Trial Eligibility Challenge (Table form of. **Figure 2**).

|  | Data | Parameters |  | Balanced Accuracy | Micro-F1 |
| --- | --- | --- | --- | --- | --- |
|  |  | Temperature | Top_p |  |  |
| 8B | Train | 0 | 1 | 0.690 | 0.842 |
|  |  | 1 | 0.5 | 0.681 | 0.810 |
|  | Test | 0 | 1 | 0.735 | 0.847 |
|  |  | 1 | 0.5 | 0.737 | 0.819 |
| 70B | Train | 0 | 1 | **0.814** | **0.901** |
|  |  | 1 | 0.5 | **0.815** | **0.897** |
|  | Test | 0 | 1 | **0.840** | **0.886** |
|  |  | 1 | 0.5 | **0.835** | **0.881** |
| Fine-tuned 8B-H-25k | Train | 0 | 1 | 0.737 | 0.872 |
|  |  | 1 | 0.5 | 0.720 | 0.864 |
|  | Test | 0 | 1 | 0.756 | 0.875 |
|  |  | 1 | 0.5 | 0.750 | 0.881 |
| Fine-tuned 8B-All | Train | 0 | 1 | 0.740 | 0.880 |
|  |  | 1 | 0.5 | 0.720 | 0.883 |
|  | Test | 0 | 1 | 0.760 | 0.874 |
|  |  | 1 | 0.5 | 0.745 | 0.878 |
| Fine-tuned<br>8B-NB-Only | Train | 0 | 1 | 0.681 | 0.828 |
|  |  | 1 | 0.5 | 0.680 | 0.832 |
|  | Test | 0 | 1 | 0.703 | 0.836 |
|  |  | 1 | 0.5 | 0.680 | 0.832 |
| Fine-tuned<br>8B-No-S | Train | 0 | 1 | 0.632 | 0.831 |
|  |  | 1 | 0.5 | 0.626 | 0.825 |
|  | Test | 0 | 1 | 0.683 | 0.809 |
|  |  | 1 | 0.5 | 0.674 | 0.800 |

**Supplementary Table 4.** Apixaban annotated data summary. There were 23 questions (15 boolean, 8 numeric) answered per patient, so for a total of 100 patients there were 2300 questions. Since there are 100 patients, the count of each answer for each question is the same number as the percentage.

Boolean
|  | Question | Answer | Count (%) |
| --- | --- | --- | --- |
| 1 | Does the note describe the patient as having **atrial fibrillation (afib)**? Answer "No" if the note describes the patient as having afib secondary to another reversible cause. | Yes | 71 (71%) |
|  |  | No | 29 (29%) |
| 2 | Does the note describe the patient as ever being diagnosed with **depression** or major depressive disorder (MDD)? Answer "No" unless the note describes a diagnosis or history of depression. | Yes | 23 (23%) |
|  |  | No | 77 (77%) |
| 3 | Does the note describe the patient as ever being diagnosed with **schizophrenia or any schizoaffective disorders**? Answer "No" unless the note describes a diagnosis or history of a schizoaffective disorder. | Yes | 2 (2%) |
|  |  | No | 98 (98%) |
| 4 | Does the note describe the patient as ever being diagnosed with **bipolar disorder**? Answer "No" unless the note describes a diagnosis or history of bipolar disorder. | Yes | 5 (5%) |
|  |  | No | 95 (95%) |
| 5 | Does the note describe the patient as ever having any **hemorrhagic tendencies** or **blood dyscrasias**? Answer "No" unless the note describes a diagnosis or history of hemorrhagic tendencies or blood dyscrasias. | Yes | 18 (18%) |
|  |  | No | 82 (82%) |
| 6 | Does the note describe the patient as having a **stroke** during this admission or within the last month? (Answer "Yes" for any recent stroke if the date is unclear, answer "No" if no stroke is mentioned or a prior stroke occurred but it was not recent) | Yes | 16 (16%) |
|  |  | No | 84 (84%) |
| 7 | Does the note describe the patient as ever having **peptic ulcer disease**? | Yes | 6 (6%) |
|  |  | No | 94 (94%) |
| 8 | Does the note describe the patient as having **serious bleeding** in the past 6 months? Answer "No" unless the note describes a serious recent bleeding issue. | Yes | 20 (20%) |
|  |  | No | 80 (80%) |
| 9 | Does the note describe the patient as having a planned or past **ablation procedure for afib**? Answer "No" unless the note includes information about a past or planned ablation for afib. | Yes | 5 (5%) |
|  |  | No | 95 (95%) |
| 10 | Does the note describe the patient as ever having valvular disease (stenosis) requiring surgery? Answer "No" if there is mention of stenosis without surgery. | Yes | 10 (10%) |
|  |  | No | 90 (90%) |
| 11 | Does the note describe the patient as having **heart failure**? | Yes | 53 (53%) |
|  |  | No | 47 (47%) |
| 12 | Does the note describe the patient as having diabetes mellitus (DM1, DM2, T2D, T1DM, T2DM)? | Yes | 44 (44%) |
|  |  | No | 56 (56%) |
| 13 | Does the note describe the patient as having **arterial hypertension** (high bp e.g. >140, or HTN)? This includes pre-existing hypertension and treated hypertension. | Yes | 82 (82%) |
|  |  | No | 47 (47%) |
| 14 | Does the note describe the patient as ever having a **stroke** or **transient ischemic attack (TIA)**? Answer "No" unless the note includes information about the patient having a prior stroke or TIA | Yes | 19 (19%) |
|  |  | No | 81 (81%) |
| 15 | Does the note describe the patient as being **unable to make medical decisions** upon discharge? Answer "No" unless there is evidence the patient cannot make their own medical decisions. Answer "Yes" if there is clear mention of dementia or the patient is deceased. | Yes | 13 (13%) |
|  |  | No | 87 (87%) |

*Numeric*
|  | Question | Mean value | Median value | Standard deviation | Range | NAs |
| --- | --- | --- | --- | --- | --- | --- |
| 1 | What is the lowest **platelet count (PLT)** mentioned in the note? Answer "NA" if no platelet count (PLT) is available in the note. | 148.53 | 147.50 | 90.8 | 15-364 | 60 (60%) |
| 2 | What is the highest **total bilirubin (TotBili, Bili)** mentioned in the note? Answer "NA" if no bilirubin value is available in the note. | 0.903 | 0.600 | 1.11 | 0.2-6.8 | 33 (33%) |
| 3 | What is the highest **aspartate aminotransferase level (AST)** mentioned in the note? Answer "NA" if no AST value is available in the note. | 194.4 | 36.0 | 1049.597 | 8-8627 | 33 (33%) |
| 4 | What is the highest **serum creatinine (Creat)** mentioned in the note? Answer "NA" if no creatinine value is available in the note. | 1.586 | 1.200 | 1.199 | 0.5-7.8 | 3 (3%) |
| 5 | What is the lowest **hemoglobin (HGB)** mentioned in the note? Answer "NA" if no HGB value is available in the note. | 10.21 | 10.15 | 2.054 | 6.0-15.9 | 2 (2%) |
| 6 | What is the highest **CHADS2** score mentioned? Answer "NA" if no CHADS2 score is in the note. | 3.95 | 3.50 | 1.39 | 1-6 | 80 (80%) |
| 7 | What is the lowest **left ventricular ejection** (LVEF, ef, ejection fraction) fraction mentioned in the note?<br>Answer "NA" if no LVEF is in the note, Answer 55 if the lowest value is 55%% or greater. | 47.89 | 50.00 | 14.4 | 20-75 | 53 (53%) |
| 8 | What is the highest **blood glucose** lab mentioned?<br>Answer "NA" if no blood glucose score is in the note. | 142.1 | 126.0 | 52.2 | 78-412 | 3 (3%) |

**Supplementary Table 5.** Hourly GPU rates. This table contains posted hourly rates for 8 x A100 GPU instances in the Eastern US region, rounded to the nearest cent from 3 major providers. The exact hardware specification is found in the Instance column. Websites were available as of September 27 2024, and are available on the Internet Archive.

| Service | Provider | Instance | Price / hour | Source |
| --- | --- | --- | --- | --- |
| **Azure** | Microsoft | ND96asr A100 v4 | \$27.20 | https://azure.microsoft.com/en-us/pricing/details/machine-learning/ |
| **cloudML** | Google | a2-highgpu-8g | \$29.39 | https://cloud.google.com/compute/all-pricing |
| **AWS** | Amazon | p4d.24xlarge | \$32.77 | https://aws.amazon.com/ec2/instance-types/p4/ |

**Supplementary Table 6.** Example generated questions by type. The following table contains 3 example questions for each question type.

| Type | Question |
| --- | --- |
| **boolean** | Does the patient have a history of septic thrombophlebitis? |
|  | Was the patient's oxygen saturation below 90% upon admission? |
|  | Is the patient's hemoglobin level within the normal range? |
| **numeric** | What was the patient's age at admission? |
|  | What was the patient's highest recorded WBC count? |
|  | What was the patient's INR on 2181-5-21? |
| **na-boolean** | Is the patient's anemia related to a chronic disease? |
|  | Is the patient a current smoker? |
|  | Has the patient undergone any prior surgery on the right upper extremity? |
| na-numeric | What is the patient's peak oxygen consumption during a cardiopulmonary exercise test? |
|  | What is the patient's estimated glomerular filtration rate (eGFR) upon admission? |
|  | What is the patient's 6-minute walk test distance? |

